# Identification of novel genetic variants in the malaria vaccine candidate PfRh5: structure-guided insights into potential function

**DOI:** 10.1101/2022.03.17.22271950

**Authors:** Khadidiatou Mangou, Adam J. Moore, Laty Gaye Thiam, Aboubacar Ba, Alessandra Orfanó, Ife Desamours, Duncan Ndungu Ndegwa, Justin Goodwin, Yicheng Guo, Zizhang Sheng, Saurabh D. Patel, Fatoumata Diallo, Seynabou D. Sene, Mariama N. Pouye, Awa Thioub Faye, Vanessa Nunez, Cheikh Tidiane Diagne, Bacary Djilocalisse Sadio, Lawrence Shapiro, Ousmane Faye, Alassane Mbengue, Amy K. Bei

## Abstract

The recent stall in the global reduction of malaria deaths has made the development of a highly effective vaccine essential. A major challenge to developing an efficacious vaccine is the extensive diversity of *Plasmodium falciparum* antigens. While genetic diversity plays a major role in immune evasion and is a barrier to the development of both natural and vaccine-induced protective immunity, it has been under-prioritized in the evaluation of malaria vaccine candidates. This study uses genomic approaches to evaluate genetic diversity in next generation malaria vaccine candidate PfRh5. We used targeted deep amplicon sequencing to identify non-synonymous Single Nucleotide Polymorphisms (SNPs) in PfRH5 (Reticulocyte-Binding Protein Homologue 5) in 189 *P. falciparum* positive samples from Southern Senegal and identified 74 novel SNPs. We evaluated the population prevalence of these SNPs as well as the frequency in individual samples and found that only a single SNP, C203Y, was present at every site. Many SNPs were unique to the individual sampled, with over 90% of SNPs being found in just one infected individual. In addition to population prevalence, we assessed individual level SNP frequencies which revealed that some SNPs were dominant (frequency of greater than 25% in a polygenomic sample) whereas most were rare, present at 2% or less of total reads mapped to the reference at the given position. Structural modeling uncovered 3 novel SNPs occurring under epitopes bound by inhibitory monoclonal antibodies, potentially impacting immune evasion, while other SNPs were predicted to impact PfRh5 structure or interactions with the receptor or binding partners. Our data demonstrate that PfRh5 exhibits greater genetic diversity than previously described, with the caveat that most of the uncovered SNPs are at a low overall frequency in the individual and prevalence in the population. The structural studies reveal that novel SNPs could have functional implications on PfRh5 receptor binding, complex formation, or immune evasion, supporting continued efforts to validate PfRh5 as an effective malaria vaccine target and development of a PfRh5 vaccine.

## Introduction

Malaria remains one of the most prevalent and deadly diseases in the world; responsible for an estimated 241 million cases of disease and 627,000 deaths annually^1^. Over 95% of these cases and deaths are attributed to the parasite *Plasmodium falciparum*. While the field has made great strides in malaria control continued advancement has been near a standstill in recent years^1^ and has even reversed course. A key advancement in *P. falciparum* control came about with the development and licensure of the RTS,S vaccine; the first malaria, and human parasite, vaccine to have ever successfully completed Phase III clinical trials and been licensed for use^2^. In the Fall of 2021, the World Health Organization officially recommended the RTS,S vaccine, for children in areas with moderate to high transmission. However, while a tremendous achievement for the field, the RTS,S vaccine shows limited efficacy that wanes with time^3,4^. One of the challenges faced by the RTS,S vaccine, and other vaccines that did not make it through the development pipeline, is genetic diversity in the target antigen^5^.

*P. falciparum*’s highly complex life cycle presents a number of potential stages to design a vaccine around^6^. The human blood-stage infection presents an Achilles’ heel in that successful invasion of human erythrocytes is essential for the completion of the parasite’s life cycle in the human host, and ultimately, transmission to the mosquito^7^. Due to the crucial nature of this life stage, the parasite has evolved a plethora of invasion ligands, which are both polymorphic and variantly expressed, that it can implement to successfully invade an erythrocyte^8^. Among these ligands is one that has been shown to be essential, and thus an attractive vaccine candidate: Plasmodium falciparum reticulocyte binding protein homologous 5 (PfRh5)^9^. PfRh5 binds to the Basigin (BSG) receptor on the erythrocyte, marking a crucial and irreversible step in the invasion process^10^. Antibodies targeting the PfRh5-BSG invasion pathway, both on the parasite (PfRh5) and human (BSG) side, have been shown to substantially inhibit invasion efficiency^9,11^. Additionally, a PfRh5 vaccine has induced neutralizing antibodies in *Aoutus* monkeys that are strain-transcendent when challenged with genetically diverse *P. falciparum* isolates *in vivo*^12,13^.

The genetic diversity that has thwarted the efficacy of previous malaria vaccines is traditionally not specifically evaluated until well into clinical trials^2^. Previous research has shown PfRh5 to be relatively conserved, with 43 non-synonymous polymorphisms (NS SNPs) documented between published and unpublished data^14–19^. Of these documented SNPs, only five have been shown to be present at frequencies above 10% in any given population, and only one (S197Y) has been shown to contribute to immune evasion from a neutralizing monoclonal antibody^20,21^. However, the sampling for this data is largely taken from lab strains and field isolates that have been adapted to *in vitro* culture. To get ahead of the curve and determine whether a PfRh5 vaccine is likely to face the challenges of natural selection, more investigation and characterization of PfRh5 genetic diversity must be done in highly endemic regions, where genetic diversity and novel SNPs are more likely to naturally occur. In this work, we seek to do just that. We sampled from active cases in Kédougou, Senegal, a highly endemic region already known to contain unique genotypes^16^.

## Results

### Characteristics of study participants

Samples used in this study were collected from Kédougou (Figure 1) in the Southeastern region of Senegal, with a high seasonal malaria transmission from May to November, annually (Ref; PNLP). Participants for this study (N = 189) were individuals aged between 2 to 72 years, presenting symptomatic P. falciparum infection, recruited from Bandafassi (25), Bantaco (24), Camp Militaire (63), Dalaba (24), Mako (43) and Tomboronkoto (10) (Figure 1B). The demographic and parasitological characteristics of the study participants are summarized in Table 1 1. There was no significant difference observed in the mean ages between the four sampling sites (ANOVA, p=0.2131), while a significant difference was observed concerning the sex ratio across sites (Chi-square, p=0.0055). Taken individually, the sex ratio was neutral in Mako and Dalaba; while that of Bantaco and Camp Militaire was in favor of males (3.8 and 2.7, respectively), and the opposite trend was observed in Bandafassi and Mako (0.8 each). No significant difference was observed in the multiplicity of infections (MOI) across sites, while the overall MOI was 4.59.

**Table 1.** Patient demographic data and Multiplicity of Infection a - indicates test performed using Chi-square b - indicates analysis performed with One-way ANOVA BF: Bandafassi; BC: Bantaco; CM: Camp Millitaire ; DB : Dalaba ; MK : Mako ; TM : Tomboronkoto; MOI: Multiplicity of Infections

| Sites | BF | BC | CM | DB | MK | TM | Total | P-value<sup>a</sup> |
| --- | --- | --- | --- | --- | --- | --- | --- | --- |
| Patients, No. | 25 | 24 | 63 | 24 | 43 | 10 | 189 |  |
| Sex ratio (Male/Female) | 0.8 | 3.8 | 2.7 | 1 | 0.8 | 1 | 1.46 | 0.0055<sup>a</sup> |
| Age (Mean, Years)<br>[Min-Max] | 17.96<br>[2-38] | 16.54<br>[2-31] | 21.27<br>[2-72] | 18.57<br>[8-31] | 18.83<br>[5-45] | 26.40<br>[12-43] | 17.76<br>[2-72] | 0.1732<sup>b</sup> |
| MOI (Mean)<br>[Min-Max] | 5.56<br>[1-11] | 5.53<br>[1-15] | 4.98<br>[1-12] | 4.31<br>[1-10] | 4.46<br>[1-12] | 4.67<br>[3-5] | 4.59<br>[1-15] | 0.0674<sup>b</sup> |

**Figure 1.**
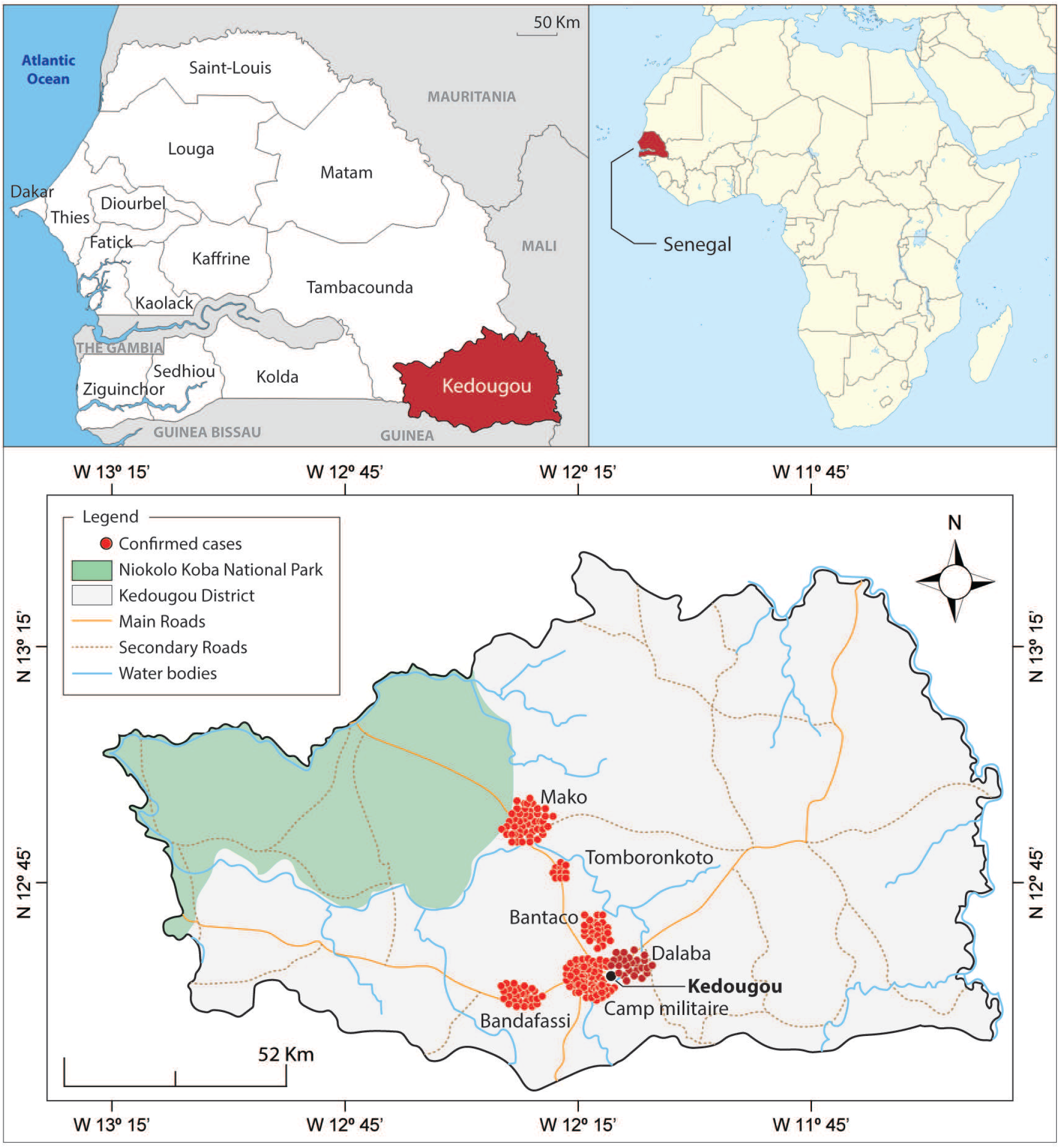
Map of the Study Population: diverse malaria epidemiology within Kédougou. Kédougou is a region in the South East of Senegal with season malaria transmission from June to December. The bottom panel displays the six sites of enrollment in the present study: Bandafassi, Bantaco, Camp Militaire, Dalaba, Mako, and Tomboronkoto. Each site has a unique epidemiology ranging from rural village (Bandafassi), to mining proximal (Bantaco), to peri-urban (Mako, Tomboronkoto, Dalaba, Camp Militaire). Red dots represent individual samples enrolled at each site.

### Overall Prevalence of known and novel SNPs in Kédougou

We undertook a population-based genetic diversity using deep amplicon sequencing data of PfRh5 obtained from 189 *P. falciparum* clinical samples using Illumina next generation sequencing. We used a very sensitive cut-off (1% variant frequency) to allow maximum SNP discovery, while applying quantitative and qualitative metrics to ensure SNP validity. Overall, 158/189 (83.59%) of the isolates contained at least one mutant allele of PfRh5, relative to the 3D7 reference genome (Figure 2A, Supplementary Figure 1). In total, 90 non-synonymous SNPs were identified from these sequences, of which, only 8 were previously reported in other studies^14,17–19^, and an additional 8 were identified in our previously published data^16^. Of the 74 novel SNPs reported here for the first time, 6 were previously described at the same position but we observed different amino acid substitutions (D52G, D127G, D379G, E362A, T384A, and Y147C) (Figure 2A); Supplementary Table 1). We include the 31 samples in our previously published genotype-phenotype association study^16^ among the 189 total here to allow for a more complete assessment of population prevalence of SNPs in PfRh5. Of the 90 SNPs reported here, 8 were present in more than one isolate, while the bulk 82/90 (91%) were singletons (detected in only individual samples) (Figure 1). The majority of the SNPs reported in this study were rare variants, as only one was detected at a prevalence greater than 3%, while only two reached over 5% prevalence (C203Y, 52.86% and I407V, 7.14%). Of the novel SNPs detected in the study, only four reach a prevalence of at least 1% in the total population - H148R (1.43%); K223R (1.43%); E306G (1.07%); and Y252H (1.07%). – and six SNPs were detected at a prevalence of 0.71% (D249G, E322G, H170R, K58E, K58R and L380P), while the remaining SNPs were all detected at a prevalence of 0.36%, corresponding to a single sample(Figure 2A).

**Figure 2.**
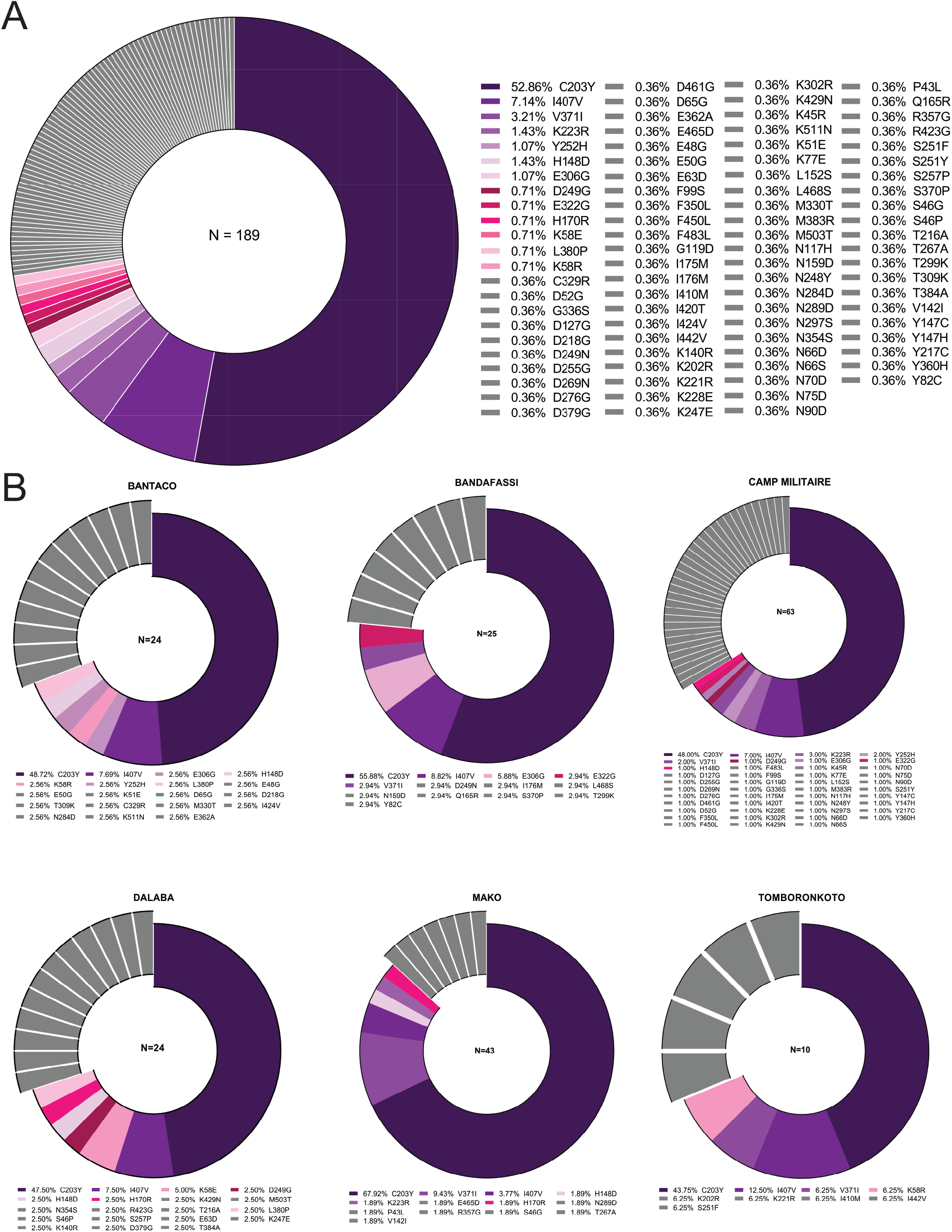
Population prevalence of known and novel SNPs in PfRh5. The prevalence of PfRh5-associated SNPs was calculated as the percentage of SNPs detected within the total sample population (A) or within site-specific sample populations (B) in Kédougou. The most prevalent SNPs, shared between two or more sites are depicted in degraded colors, while SNPs detected in individual isolates are depicted in grey (A). Exploded slices in (B) represent SNPs unique to the respective site, while the shared SNPs are shown in matched colors.

### Site Specific Prevalence of known and novel SNPs in Kédougou

The prevalence of identified SNPs were estimated for each sampling site. As seen in the overall parasite population, the bulk of the isolates reported in the present study presented a mutant allele of PfRh5. Bantaco presented the highest number of mutant alleles (91.67%) while Bandafassi, Camp Militaire, Mako and Tomboronkoto presented relatively similar prevalences of mutant alleles (80%, 80.95%, 79.17%, 83.72% and 80%, respectively) (Supplementary Figure 1B-G). Across sites, Camp Militaire presented the highest number of individual SNPs (43 SNPs), followed by Bantaco and Dalaba (19 SNPs each), while samples from Bandafassi and Mako harboured the same number of individual SNPs (13 SNPs each), and the fewest SNPs were detected in isolates from Tomboronkoto (9 SNPs). Interestingly, the bulk of the SNPs detected were unique to the individual sampling sites (not detected at another site). All SNPs unique to a site were additionally only found in a single individual. Isolates from Camp Militaire carrying the highest number of unique SNPs (34/43; 79.06%), followed by Dalaba (12/18; 66.67%), Bantaco (12/19; 63.16%) and Bandafassi (8/13; 61.54%), while the least number of unique SNPs was detected in Mako (7/13; 53.85%) and Tomboronkoto (5/9; 55.56%), roughly proportional to the overall sample numbers from each site (Figure 2B). Of the remaining SNPs, only C203Y was shared across all sites, while the I407V and V371I mutations were shared in all but one site; I407V was absent from Mako, while V371I was absent from Dalaba. The H148R mutation was observed in isolates from Bantaco, Camp Militaire, Dalaba and Mako, while E306G was only common in isolates from Bantaco, Bandafassi and Camp Militaire. Some SNPs (K58R, H170R, D249G, Y252H, E322G and L380P) were only shared between two sites (Figure 2B)).

### Frequency of novel SNPs in individual samples

The sequence reads from next generation sequencing experiments were used to assess the frequency of the individual SNPs detected in this study within an individual sample. As the bulk of the isolates analyzed here are mixed genotypes (Average MOI 4.59, ranging from 1-15 genotypes per sample (Table 1). Due to this high degree of polygenomic infections, and the difficulty with disentangling individual genomes as well as the corresponding parasite burden or parasitemia of each genome in a sample, we calculated the frequency of a variant (SNP) for each isolate based on the overall variant read frequency among reads from individual samples. This allows us to determine the percentage of variant reads among the total at a position and to average across all complex polygenomic infections in the population. Upon analysis, the individual SNPs were categorized based on read frequency within individual samples into low frequency (<2%), intermediate frequency (2-25%) and high frequency (>25%) frequency SNPs (Figure 3). Most SNPs detected high frequencies in individual isolates were also found at a high population prevalence. A few SNPs were identified at a high frequency in individual samples, but a relatively low population prevalence (in all cases observed in a single sample): F350L, N248Y, K51E, N289D, P43L and S46P with frequencies ranging from 28.1 – 47.98%; and D269N, T299K, S46G, N159D, K429N and R357G with high frequencies ranging from 60.41 – 76.37%. The majority of the novel SNPs described here are present at low to intermediate frequency within a sample and rare in the population (Figure 3).

**Figure 3.**
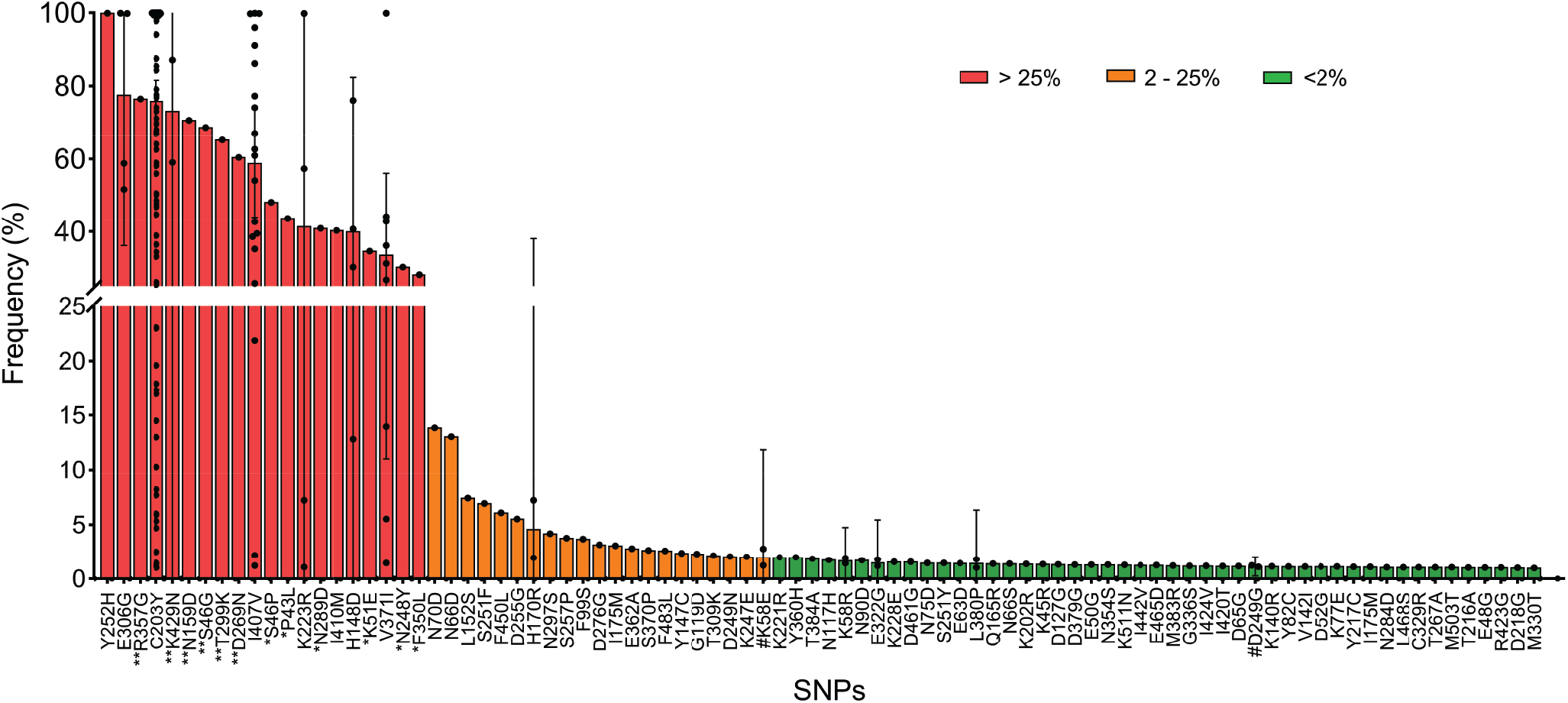
Frequency of PfRh5 SNPs in individual complex infections. SNP frequency was determined from the sequencing read data and calculated as the percentage of the variant read coverage relative to the coverage at the variant position. Using quantitative read coverage, we are able to calculate frequency within the overall *Plasmodium* genetic material in a given sample, incorporating multiple genotypes as well as variable parasitemia across genotypes. SNPs are categorized as low <2% (green), intermediate 2-25% (orange) or high frequency SNPs >25% (red), based on their respective frequency with the sample population. Additional symbols are used to highlight SNPs occurring at high prevalence but low frequency (““), SNPs occurring at low prevalence relatively high (*) or very high frequency (**).

### Structure-based insights on potential SNP function

We used structure-based approaches to thread the identified SNPs into the crystal structure of PfRh5 in complex with its receptor basigin and binding partner PfCyRPA (Figure 4A). The complex was built by superposing the RH5 of RG5-BSG complex (PDB id: 4U0Q) and RH5-CyRPA complex (PDB id: 6MPV) The structural threading analysis revealed four groups of SNPs with different predicted functional outcomes. One of the groups includes PfRh5-associated polymorphisms predicted to alter binding to the Basigin receptor (e.g. C203Y, S197Y, N354S, F350L, R357G, E362A/D) (Figure 4B-E), while another subset of SNPs was predicted to impact PfRh5 binding to its partner PfCyRPA (e.g. M503T) (Figure 4B-E). While the majority of the identified SNPs were predicted to partially alter the structure of PfRh5 (Figure 4F-J), there was a particular set of SNPs for which no structural data was available, but occurring at high frequencies within the sampled populations. Interestingly, our analysis also revealed a group of SNPs occurring under known mapped binding epitopes for inhibitory antibodies (S197Y, K202R, R357G, and E362A). Of these, only S197Y has been shown to result in immune escape in parasites harboring this mutation^21^. These remaining three SNPs are novel, with K202R having been reported in our previous study^16^, and mutations have previously been described at amino acid position E362, but with a different amino acid change (E362D)^14^ compared to E362A reported here.

**Figure 4.**
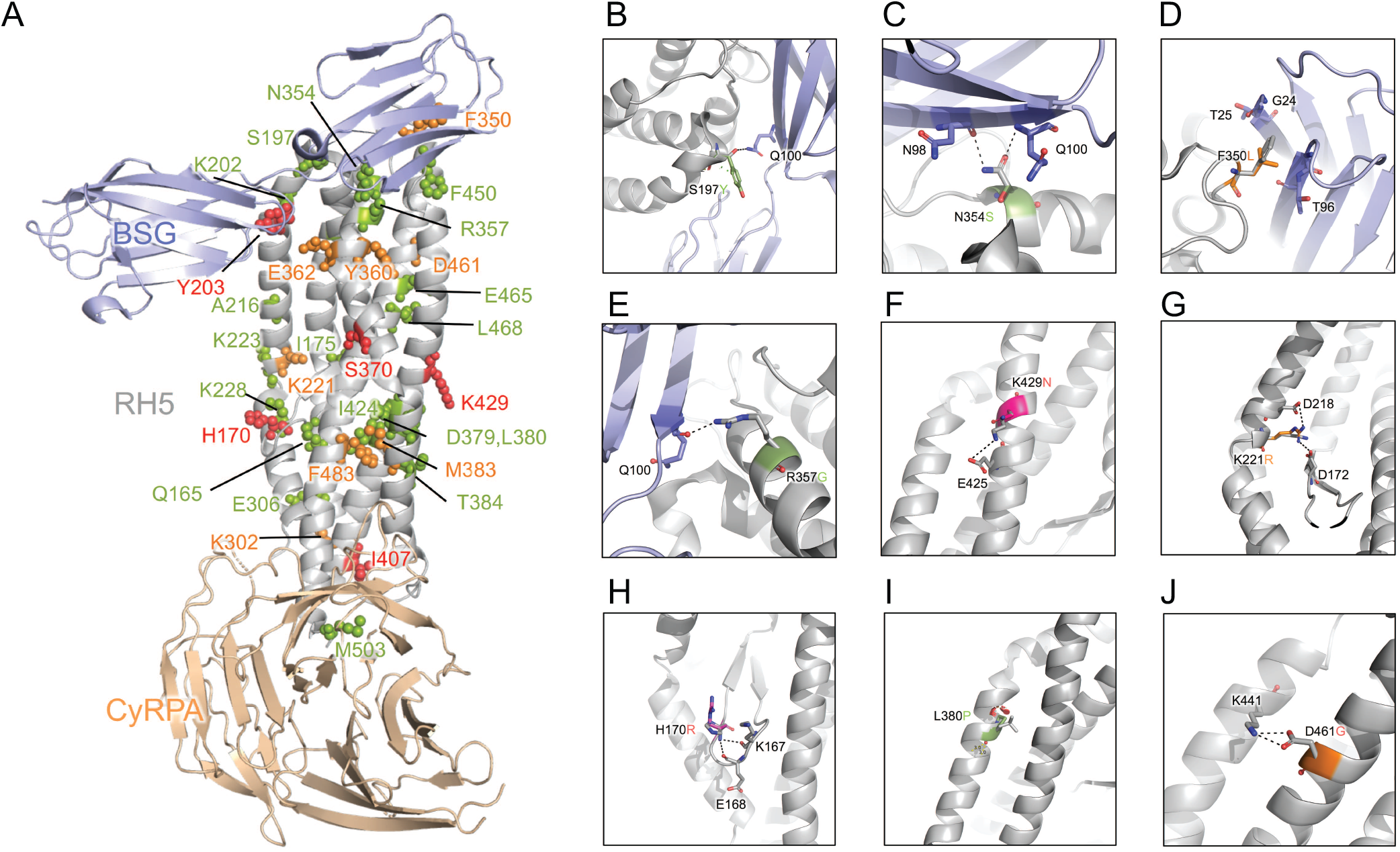
Structure-Function Predictions for novel SNPs identified in PfRh5. A. Location of SNPs in the BSG-RH5-CyRPA complex. The complex was built by superposing the RH5 of RG5-BSG complex (PDB id: 4U0Q) and RH5-CyRPA complex (PDB id: 6MPV). BSG, RH5 and CyRPA are shown by light blue, grey and light orange, respectively. B-J. Small panels highlight the predicted effect of selected SNPs, the black dotted lines represent hydrogen bonds or salt bridges. From top-left to bottom-right: SNPs that potentially impact binding of PfRh5 and BSG (B-E); C203Y, S197Y, N354S, F350L, R357G), SNPs that potentially alter the structure of PfRh5 (F-J; K429N, D461G, L380P, H170R, K221R). SNPs reaching significance above the 1% discovery threshold are color coded according to frequency in individual samples with low frequency <2% (green), intermediate frequency 2-25% (orange), high frequency >25% (red).

## Discussion

We sought to provide a thorough analysis of the genetic diversity present in the *P. falciparum* gene PfRh5 in Kédougou, Senegal, to further assess if this gene is an adequate target for a highly effective second-generation malaria vaccine. We found that PfRh5 exhibits more abundant genetic diversity in field isolates than previously thought, including the discovery of 74 novel SNPs. Implementing deep amplicon NGS on our 189 samples and setting a sensitive discovery threshold of 1% prevalence allowed us to discover these previously undescribed SNPs.

As Kédougou is a region with significant polyclonality, ranging from 1-15 genotypes per patient and an average MOI 0f 4.59, it was important to assess not just the presence of SNPs per individual, but the frequency of these SNPs in the complex mixture of parasite genomes in each sample. To this end, we assessed the “frequency” of each SNP by assessing the percentage of variant reads in the total reads mapped to a given position in PfRh5. This value allows us to calculate quantitatively the number of variant reads in the total, taking into account polygenomic infections and also varying parasitemias of each genotype in the infected individual. Using this approach, we were able to classify the SNPs into those that were high (>25%), intermediate (2-25%), and low (<2%) in samples containing these SNPs. While most SNPs that were among the most prevalent in the population were also high frequency in individual positive samples, there were some exceptions whereby a single sample contained a SNP (low population prevalence), yet that SNP was present in a high frequency of the reads. Likewise, some SNPs were present in multiple samples resulting in a higher population prevalence, but were always at a low frequency in the individual sample. These distinctions will be important to explore through functional assays assessing the fitness of individual SNPs when compared to wild-type. It is also possible that these low frequency SNPs may also represent deleterious mutations that have not yet undergone purifying selection in the host.This division into frequency gives a more detailed picture of common and rare SNPs as it incorporates the complexity of the highly mixed infections. Describing SNPs in terms of frequency is also helpful in prioritizing SNPs for downstream follow-up.

Our site-specific analysis identified varying trends across sites that can be partially explained by their unique epidemiologies, locations, and capacities. Camp Militaire is a large military clinic that has a very large catchment area. As such, it is not surprising that this site included more samples through passive case detection and identified more individual SNPs than the others. Bantaco presents an interesting situation where over 90% of samples contained at least one SNP, about 10% higher than the other sites. Bantaco is a village that contains one of the largest traditional gold mining sites in the region. This mining site attracts workers from a variety of locations, including across the borders of Mali, Gambia, Guinea-Bissau, and Guinea, as well as countries that do not share a border such as Togo and Côte d’Ivoire, among others. This unique ecology facilitates the import of unique parasite variants from the West African sub-region, resulting in the mixing of parasite populations, naturally increasing the population-level genetic diversity. The Bantaco and Bandafassi sites are more isolated than the other sites, which sit near roads and can service patients that are not necessarily currently living near the clinic. We expected to see more allele sharing across Mako, Camp Militaire, Dalaba, and Tomboronko given their proximity to roads and thus the higher likelihood of patient populations coming from similar areas. This was observed with certain SNPs, such as D249G and H170R being found in Dalaba, Mako, and Camp Militaire. C203Y was the only SNP found across all sites, which is expected given our previous documentation of its high prevalence in the region^16^ and in other regions of Senegal^17^. Overall, the sample size from each clinic does not truly permit quantitative comparisons of genetic diversity between sites; however some of these trends are interesting and would warrant further investigation in prospective targeted studies.

While characterization of genetic diversity in vaccine candidate antigens is important, ultimately functional studies will be needed to appreciate the role of specific polymorphisms on biological functions like receptor binding, PfRh5-PfCyRPA-PfRipr (RCR) complex formation, and immune evasion. To this end, to generate testable hypotheses as to which SNPs may be important to prioritize for future functional studies, crystal structure threading of the observed SNPs allowed us to uncover potential functional impacts on the PfRh5 complex, its binding partner PfCyRPA, and the associated erythrocyte receptor basigin. Most of the SNPs are predicted to result in minor changes to the protein’s structure. Importantly, our analysis found 4 SNPs (S197Y, K202R, R357G, and E362A) that occur beneath well-defined epitopes bound by neutralizing human monoclonal antibodies from PfRh5 vaccinated individuals^21^. Of these, only S197Y has been previously described and was found to aid parasites in evading the immune response^21^. Lack of crystal structure data for a subset of our observed SNPs prevented structural threading, thus we cannot predict the potential functional impact of these SNPs.

Our study has some limitations that are important to note. First, the sample sizes across sites are not equivalent due to enrollment through passive case detection of febrile patients who come to the respective clinic and give informed consent to participate in the study after testing positive for *P. falciparum*. This sampling strategy does not represent a true cross-sectional population prevalence as might be achieved using systematic cross sectional sampling or genotyping from discarded diagnostic materials. This could potentially bias analysis of genetic diversity towards parasite strains that may cause more prominent clinical disease that influences the patient to seek care. Future work can address this by sampling across the clinical presentation spectrum, including active surveillance of asymptomatic cases.

While our structural modeling can imply potential functional outcomes of the SNPs identified in PfRh5, further experimental work is needed to fully assess the potential functional impacts of these SNPs, in binding to the Basigin receptor, binding to PfRh5 binding partners, and in immune evasion in the presence of naturally acquired and vaccine-induced immune responses. This study will inform downstream biochemical and functional genetic approaches to evaluate the role of each SNP in receptor binding, invasion, and immune evasion.

## Methods

### Study Sites

This study was conducted with ethical approval from the National Ethics Committee of Senegal (CNERS) and the Institutional Review Board of the Yale School of Public Health. All research was performed in accordance with relevant guidelines and regulations, and informed consent was obtained from all participants and/or their legal guardians.

Samples used in this study were collected as part of ongoing surveillance conducted by Institut Pasteur de Dakar investigating causes of febrile illness. Patients were recruited from six health posts across Kédougou, Senegal: Bantaco, Mako, Camp Militaire, Bandafassi, Dalaba, and Tomboronkoto. The only eligibility criteria for the main study was the presence of a fever (temperature greater than or equal to 38 degrees C) and/or a fever in the past 24 hours. Study clinical staff assessed eligibility and obtained informed consent from patients who tested positive for *P. falciparum* on a Pf-specific HRP2/3 rapid diagnostic test (RDT). After a venous blood draw of 5ml in an EDTA vacutainer was obtained from consenting, enrolled patients, samples transported at room temperature to the field lab for processing; no more than 6 hours between draw and processing. Thin and thick blood smears were made for each sample to confirm infection with only *P. falciparum* by microscopy.

### DNA Extraction, Amplification, and Sequencing

DNA was extracted from dried blood spots (DBS) using QIAmp DNA Blood Mini Kit according to manufacturer’s instructions. PfRh5 exon 2 was PCR amplified using previously described primers^17^ and high fidelity polymerase. PCR amplicon size and purity were confirmed on an agarose gel prior to Next Generation Sequencing (NGS). PCR amplicons for PfRh5 exon 2 were bead-purified (Omega) and quantified by Qubit and adjusted to equivalent concentration. Library prep was performed with Nextera XT using unique dual indexes (UDIs). After library prep, samples were again bead-purified and quantified by qPCR using Roche KAPA Library Quantification Kit. All samples were normalized to a concentration of 4nM. After this normalization, quantified and normalized libraries were pooled into 8-sub pools. These 8 sub-pools were bead-purified and quantity was measured by KAPA qPCR. The 8 sub-pools were normalized and combined in equal parts to form one final pool. This final pool was sent to the Yale Center for Genome Analysis (YCGA) for sequencing on an Illumina NovaSeq 6000 platform with targeted coverage of 500,000 reads per sample.

De-multiplexed forward and reverse sequencing reads were obtained for each sample. These reads were imported to Geneious individually then paired. This was followed by trimming using the Geneious plugin BBDuk. A minimum Quality Score (Q) of 30 was set for the trimming with a minimum length of 75 base pairs, as we were expecting reads around 100 base pairs. The trimmed sequences were mapped against a 3D7 reference genome (PF3D7_ 0424100) that had been annotated with all known synonymous and non-synonymous mutations. The mapping was set for two iterations and coverage criteria was set at 1000 reads. The criteria for SNP calling was set to a minimum frequency of 0.01 (1%) and 1000 read coverage. Sequence data and SNP analysis was performed by at least 3 individuals for each sample.

### Multiplicity of Infection

Multiplicity of infection (MOI) for each sample was determined through genotyping the merozoite surface proteins (MSP) with a nested PCR on DNA extracted from whole blood, as described by Snounou^22^.

### Structure modeling of SNPs

The structures of BSG-RH5 and CyRPA-RH5 were downloaded from the Protein Data Bank (PDB, https://www.rcsb.org/) under accession number 4U0Q and 6MPV, respectively. The BSG-RH5-CyRPA complex was built by superposing the RH5 in the BSG-RH5 and CyRPA-RH5 complexes. Pymol version 2.3.2 was used to predict the effect and to plot the structural location of each SNP^23^.

### Statistical Analysis

Summary statistics of patient demographics were performed and comparisons were made across the six study sites. Sex ratios were compared using the Chi-squared test across sites. Comparisons of average age and MOI across sites were compared using One-way ANOVA. P-values 0.05 are considered significant.

## Data Availability

Sequencing Reads associated with this study have been deposited in the NCBI SRA with the BioProject Accession: PRJNA811159.
All data produced are available online at https://www.ncbi.nlm.nih.gov//bioproject/PRJNA811159

## Acknowledgements

We would like to thank Lt. Dr Chares Latyr Ndiaye and Lamine Kane from Camp Militaire, Moctar Mansaly and Gerald Keita from Bandafassi, Safietou Sane and Astou Ndiaye from Mako, Adama Gueye from Bantaco, Souleymane Ngom from Dalaba, Der Ciss from Tomboronkoto and all the healthcare workers at these sites for their partnership with Institut Pasteur Dakar. We would also like to thank the people of Kédougou for their invaluable contributions to this work. We would like to thank the Yale Center for Genome Analysis (YCGA). This publication refers to data generated by the Pf3k project (www.malariagen.net/pf3k), representing 3,248 samples from 40 separate locations in 20 countries, and data published by Manske and colleagues^14^.

## Author contributions statement

A.K.B. conceived the experiments. A.K.B., A.M., L.S., and O.F. supervised the research. K.M., F.D., S.D., M.N.P., B.D.S. collected the samples. B.D.S. and C.T.D. assisted with geolocation. A.J.M., K.M., A.T.F, A.B., D.N.N., J.G., F.D., S.D., and M.N.P. conducted the experiments. Y.G., Z.S., S.D.P performed structure modeling. A.J.M., L.G.T, K.M., A.B, V.N. and A.K.B. analyzed the results. A.J.M., L.G.T., K.M., and A.K.B. wrote the manuscript. All authors reviewed the final manuscript.

## Additional information

### Data Accessibility

Sequencing Reads associated with this study have been deposited in the NCBI SRA with the BioProject Accession: PR-JNA811159.

## References

1. World malaria report 2021. Report, World Health Organization (2021).

2. Draper, S. J. et al. Malaria Vaccines: Recent Advances and New Horizons. Cell Host & Microbe 24, 43–56, DOI: 10.1016/j.chom.2018.06.008 (2018).

3. Olotu, A. et al. Seven-Year Efficacy of RTS,S/AS01 Malaria Vaccine among Young African Children. New Engl. J. Medicine 374, 2519–2529, DOI: 10.1056/NEJMoa1515257 (2016).

4. RTS,S Clinical Trials Partnership. Efficacy and safety of RTS,S/AS01 malaria vaccine with or without a booster dose in infants and children in Africa: final results of a phase 3, individually randomised, controlled trial. Lancet (London, England) 386, 31–45, DOI: 10.1016/S0140-6736(15)60721-8 (2015).

5. Neafsey, D. E. et al. Genetic diversity and protective efficacy of the rts,s/as01 malaria vaccine. N Engl J Med 373, 2025–37, DOI: 10.1056/NEJMoa1505819 (2015).

6. Cowman, A. F., Healer, J., Marapana, D. & Marsh, K. Malaria: Biology and Disease. Cell 167, 610–624, DOI: 10.1016/j.cell.2016.07.055 (2016).

7. Patarroyo, M. E., Alba, M. P., Reyes, C., Rojas-Luna, R. & Patarroyo, M. A. The Malaria Parasite’s Achilles’ Heel: Functionally-relevant Invasion Structures. Curr. Issues Mol. Biol. DOI: 10.21775/cimb.018.011 (2016).

8. Wright, G. J. & Rayner, J. C. Plasmodium falciparum Erythrocyte Invasion: Combining Function with Immune Evasion. PLOS Pathog. 10, e1003943, DOI: 10.1371/journal.ppat.1003943 (2014).

9. Crosnier, C. et al. Basigin is a receptor essential for erythrocyte invasion by Plasmodium falciparum. Nature 480, 534 (2011).

10. Cowman, A. F. et al. Functional analysis of proteins involved in Plasmodium falciparum merozoite invasion of red blood cells. FEBS Lett. 476, 84–88, DOI: 10.1016/S0014-5793(00)01703-8 (2000).

11. Douglas, A. D. et al. Neutralization of Plasmodium falciparum Merozoites by Antibodies against PfRH5. The J. Immunol. 192, 245–258, DOI: 10.4049/jimmunol.1302045 (2014).

12. Douglas, A. D. et al. The blood-stage malaria antigen PfRH5 is susceptible to vaccine-inducible cross-strain neutralizing antibody. Nat. Commun. 2, 601, DOI: 10.1038/ncomms1615 (2011).

13. Douglas, A. et al. A PfRH5-Based Vaccine Is Efficacious against Heterologous Strain Blood-Stage Plasmodium falciparum Infection in Aotus Monkeys. Cell Host & Microbe 17, 130–139, DOI: 10.1016/j.chom.2014.11.017 (2015).

14. Manske, M. et al. Analysis of Plasmodium falciparum diversity in natural infections by deep sequencing. Nature 487, 375–379, DOI: 10.1038/nature11174 (2012).

15. Pearson, R. D., Amato, R., Kwiatkowski, D. P. & Project, M. P. f. C. An open dataset of Plasmodium falciparum genome variation in 7,000 worldwide samples. bioRxiv 824730, DOI: 10.1101/824730 (2019).

16. Moore, A. J. et al. Assessing the functional impact of pfrh5 genetic diversity on ex vivo erythrocyte invasion inhibition. Sci Rep 11, 2225, DOI: 10.1038/s41598-021-81711-9 (2021).

17. Patel, S. D. et al. Plasmodium falciparum Merozoite Surface Antigen, PfRH5, Elicits Detectable Levels of Invasion-Inhibiting Antibodies in Humans. The J. Infect. Dis. 208, 1679–1687, DOI: 10.1093/infdis/jit385 (2013).

18. Ajibaye, O. et al. Genetic polymorphisms in malaria vaccine candidate Plasmodium falciparum reticulocyte-binding protein homologue-5 among populations in Lagos, Nigeria. Malar. J. 19, 6, DOI: 10.1186/s12936-019-3096-0 (2020).

19. Ndwiga, L. et al. The plasmodium falciparum rh5 invasion protein complex reveals an excess of rare variant mutations. Malar J 20, 278, DOI: 10.1186/s12936-021-03815-x (2021).

20. Bustamante, L. Y. et al. A full-length recombinant Plasmodium falciparum PfRH5 protein induces inhibitory antibodies that are effective across common PfRH5 genetic variants. Vaccine 31, 373–379, DOI: 10.1016/j.vaccine.2012.10.106 (2013).

21. Alanine, D. G. et al. Human Antibodies that Slow Erythrocyte Invasion Potentiate Malaria-Neutralizing Antibodies. Cell 178, 216–228.e21, DOI: 10.1016/j.cell.2019.05.025 (2019).

22. Snounou, G. Genotyping of Plasmodium spp. Nested PCR. Methods Mol. Medicine (2002).

23. Schrödinger, LLC. The PyMOL molecular graphics system, version 1.8 (2015).

