## Supplemental Figure 1 for "Identification of novel genetic variants in the malaria vaccine candidate PfRh5: structure-guided insights into potential function"

### This PDF file includes:

Fig. S1
Table S1

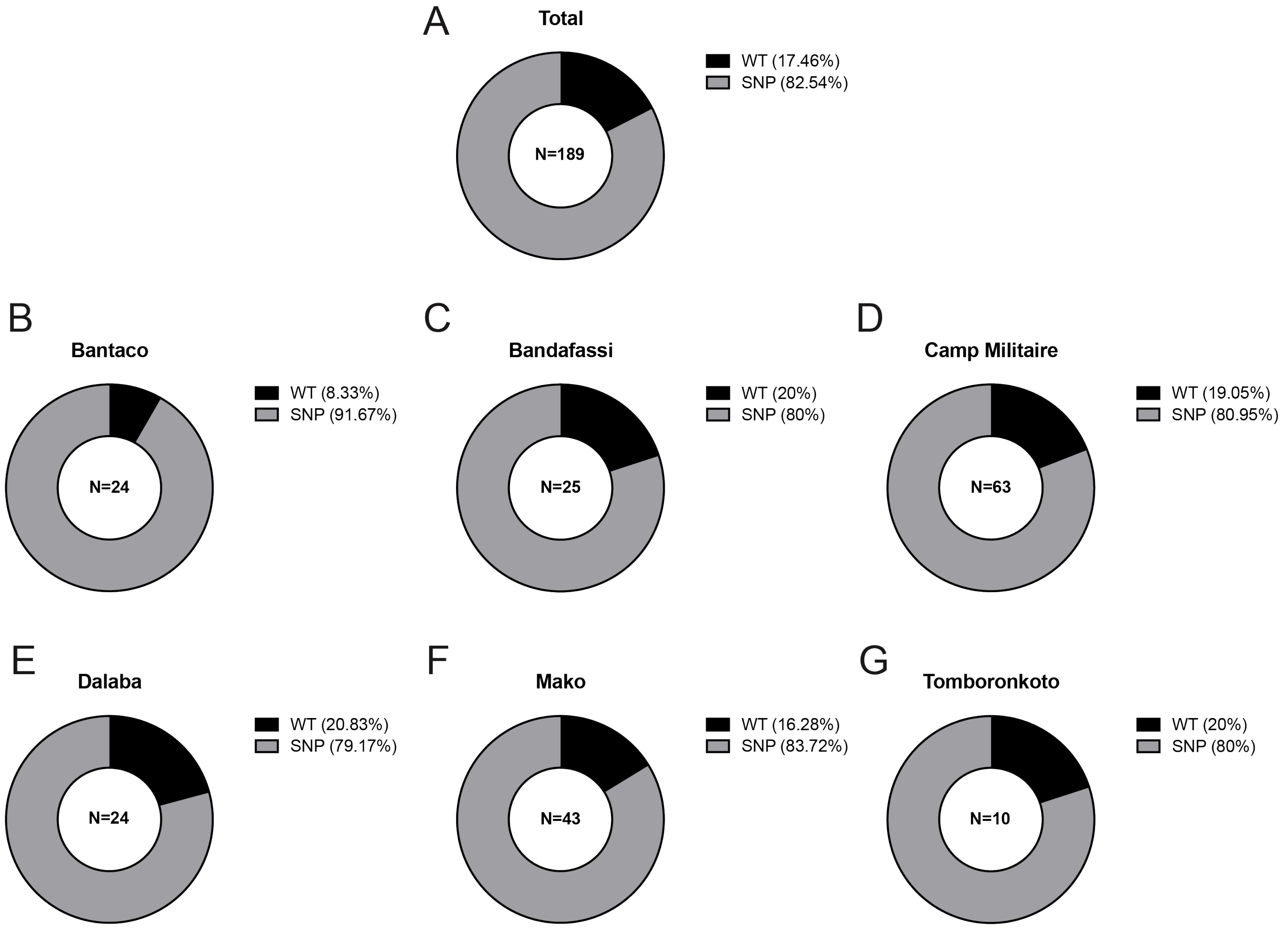


**Fig. S1. Population prevalence of wild-type and mutant PfRh5 alleles**

The prevalence of wild-type (3D7-allele) and mutant PfRh5 alleles (any mutation) was calculated as the percentage of isolates containing one or more SNPs at the discovery threshold (>1% variant frequency) relative to the total sample population (A) or site-specific sample populations (B-G) in Kédougou.

**Table S1. Description of the SNPs identified in patient samples from Kédougou**

Table S1 is included as a supplemental file (.xls). Sampling sites are abbreviated as follows: Bandafassi (BF), Bantaco (BC), Camp Militaire (CM), Dalaba (DB), Mako (MK) and Tomboronkoto (TM). For each isolate, non-synonymous amino acid substitutions are indicated along with their respective statistics (Coverage at SNP position, Variant Read coverage, and Variant Read frequency).
